# Elucidating Emotional Patterns in Autism Spectrum Disorder: BERT-Based Analysis Reveals Novel Dimensional Structure

**DOI:** 10.1101/2025.05.15.25326247

**Authors:** Muneaki Kanno, Yuka Yoshida, Momoko Fujihashi, Nana Takahashi, Takuma Numazawa, Yunosuke Mizuno

## Abstract

**Background:** Autism spectrum disorder (ASD) is associated with difficulties in emotion recognition and regulation, which complicates clinical support and treatment. While natural language processing (NLP) has enabled automated emotion analysis, few studies have investigated emotion structure in ASD using dimensional approaches.

**Objective:** To develop a BERT-based model for estimating eight-dimensional emotion profiles and to apply this model to clinical records of adolescents with ASD to elucidate characteristic affective patterns.

**Methods:** We fine-tuned five Japanese-language BERT variants using the WRIME dataset, which contains annotations for eight basic emotions with graded intensity. The best-performing model was applied to clinical records from 14 adolescents with ASD, yielding emotion profile vectors for 1,239 clinical sessions. Principal component analysis (PCA) was conducted on the resulting emotion vectors to identify dominant affective dimensions.

**Results:** The best-performing model (*tohoku-nlp/bert-large-japanese-v2*) achieved an accuracy of 78.9% and a cosine similarity of 94.1%. PCA revealed a primary emotional dimension dominated by sadness, and a secondary axis characterized by a contrast between disgust and anticipation. These patterns diverge from canonical emotion models such as Plutchik’s circumplex and suggest a distinct emotional architecture in ASD.

**Conclusion:** This study demonstrates the utility of fine-tuned BERT models in extracting nuanced emotion profiles from clinical text. The identified emotional dimensions may provide a basis for developing more personalized support strategies for individuals with ASD.

## 1. Introduction

Individuals with autism spectrum disorder (ASD) exhibit impaired recognition of all basic emotions, including happiness, sadness, anger, surprise, fear, and disgust[1]. Additionally, they often employ fewer adaptive emotion regulation strategies and more maladaptive strategies such as suppression and avoidance[2]. These difficulties in recognizing and regulating emotions create significant challenges for individuals with ASD, necessitating that medical professionals and caregivers develop a deeper understanding of the emotional characteristics associated with this condition to provide effective support.

To systematically analyze emotions, researchers have developed various frameworks. While the Ekman model has been influential, Plutchik’s Wheel of Emotions offers particularly comprehensive insights through its structure of eight basic emotions that can combine to form more complex emotional states[3]. This framework has gained widespread adoption in psychological research due to its robust structure and notably surpasses the Ekman model by including positive emotion categories such as trust and anticipation[4].

Recent advancements in natural language processing have facilitated the application of established emotion frameworks to machine learning models and large language models (LLMs). Emotion analysis, a critical task in this field, involves detecting and classifying emotions within textual data. In the medical domain, this approach has several valuable applications, such as monitoring health conditions through detecting emotions in clinical records [5] and identifying mental illnesses, including post-traumatic stress disorder, depression, and ASD[6-9].

However, most existing methods have primarily focused on sentiment analysis or basic emotion detection, which may not fully capture the nuanced complexity of human emotional experiences. We hypothesized that representing Plutchik’s eight fundamental emotions as an eight-dimension profile vector could enable a more detailed analysis of emotional states. Furthermore, we propose that adapting this approach for individuals with ASD could provide insights into emotion structures characteristic of the disorder.

To investigate these possibilities, this study has two primary objectives. First, we aim to fine-tune LLMs using an emotion-annotated Japanese dataset to develop a model optimized for eight-dimensional emotion profile analysis. Second, we seek to apply this model to clinical records of adolescent patients with ASD to elucidate the emotional characteristics associated with the disorder. Through this approach, we aim to generate insights that may contribute to the development of more effective support strategies for individuals with ASD.

## 2. Methods

### 2.1. Datasets

This study utilized the WRIME dataset, which contains 43,200 Japanese short text posts from social networking services, annotated by 80 writers and three readers[10]. The readers assessed the intensity of Plutchik’s eight basic emotions (joy, sadness, anticipation, surprise, anger, fear, disgust, and trust) [3]using a four-point scale (none, weak, medium, and strong).

The decision to employ a model trained on social media text rather than medical corpora was deliberate, as clinical psychiatric interactions primarily involve everyday language rather than technical terminology. This approach aligns with the natural communication patterns observed in clinical settings.

Precious research by Kajiwara et al. [10]demonstrated that BERT models [11] achieve superior performance when fine-tuned on WRIME using objective reader annotations rather than subjective writer data. Accordingly, we employed the averaged readers’ scores in our implementation. For model training and evaluation, we utilized the WRIME dataset partition of 40,000 posts for training, 1,200 for validation, and 2,000 annotated posts for testing.

### 2.2. Model Training

We utilized *tohoku-nlp/bert-base-japanese-v3[12], tohoku-nlp/bert-large-japanese-v2[12], nlp-waseda/roberta-large-japanese[13]. FacebookAI/xlm-roberta-large*, and *ku-nlp/deberta-v2-large-japanese* as base models, all of which were pre-trained in Japanese corpora and obtained from the Hugging Face repository distributed under the Apache License 2.0, CC-BY-SA-4.0, and MIT License. Fine-tuning was performed on the WRIME dataset, optimizing with a batch size of 32, a learning rate of 2 × 10^−5^, and a total of 4 epochs. All computations were executed on an NVIDIA A100 GPU in Google Colaboratory environment. To evaluate model performance, we assessed accuracy, weighted F1 score, and average cosine similarity with respect to eight-dimensional vector representations encoding emotional profile. For the subsequent analysis, we used the best performing model.

### 2.3. Participants

We retrospectively identified 14 adolescents (10 males and 4 females) diagnosed with ASD based on DSM-5 criteria who had their first consultation between 2011 and 2017. Their electronic health records (EHR) from the initial consultation through December 2024 were exported as UTF-8 plain text. Each consultation day was treated as a single session. Patient utterances were identified by speaker tags (“>” prefix). Texts were normalized to Unicode NFC; half-width katakana were converted to full-width; variant characters were unified. Proper names and facility identifiers were not masked. During each medical consultation, the psychiatrist recorded the session in the electronic medical record system in text format, aiming to capture the content as accurately as possible. To compile the text dataset for further analysis, we extracted participants’ utterances from these text-based records on a daily basis. All analyses were performed on an offline Mac mini (Apple M1, macOS Sonoma 14.4) with access restricted to the first author only (Touch ID plus a strong password). After extraction, the clinical records were kept as plain text—including all proper names—and fed directly into the BERT-based model in inference-only mode (model.eval(); with torch.no_grad()), ensuring that no parameter updates were made at any point.

### 2.4. Feature Extraction

We extracted emotion profile estimates from a text dataset of participant sessions using our best-performing model. This model generated intensity estimates (ranging from 0 to 1) for each of Plutchik’s basic emotions through its final softmax output layer.

### 2.5. Statistical Analyses

We performed principal component analysis (PCA) on the emotion profile estimation task. Since these estimates were generated from the softmax output of the final layer, they inherently summed to one for each session. We deliberately chose to use these raw softmax outputs without compositional data analysis techniques (such as centered log-ratio transformation) because the relative frequencies of emotions carry meaningful clinical information in our context. This approach preserves both the compositional relationship between emotions and their absolute prevalence, which is particularly relevant when studying emotion profiles in clinical populations.

No additional standardization was applied before performing PCA. The number of principal components to retains was determined based on the cumulative explained variance criterion. The first four components were selected, accounting for approximately 90% of the variance. We performed PCA in Python (version 3.10.12), using the scikit-learn library (version 1.2.1).

### 2.6. Ethics

This study was approved by the Ethical Review Committee of the Yamagata University Faculty of Medicine (No. 2023-159). All participants provided informed consent, either by signing a written consent form or choosing to opting out.

## 3. Results

### 3.1. Model Performance

The performance of the models was evaluated on emotion classification and emotion profile estimation using three metrics: accuracy, weighted F1 score, and cosine similarity, as shown in Table 1. The best-performing model on the test dataset was based on *tohoku-nlp/bert-large-japanese-v2*, achieving an accuracy of 78.9%, a weighted F1 score of 78.4%, and an average cosine similarity of 94.1%. The confusion matrix is shown as Figure 1.

**Table 1.** Evaluation of the models after fine-tuning Base model Accuracy.

| Base model | Accuracy | Weighted F1 score | Average cosine similarity |
| --- | --- | --- | --- |
| *tohoku-nlp/bert-large-japanese-v2<sup>1)</sup>* | 78.9% | 78.4% | 94.1% |
| *nlp-waseda/roberta-large-japanese<sup>2)</sup>* | 76.0% | 75.7% | 93.6% |
| *FacebookAI/xlm-roberta-large<sup>3)</sup>* | 73.3% | 72.6% | 93.5% |
| *ku-nlp/deberta-v2-large-japanese<sup>2)</sup>* | 76.4% | 75.7% | 93.4% |
| *tohoku-nlp/bert-base-japanese-v3<sup>1)</sup>* | 74.9% | 73.4% | 93.2% |

1) Apache License 2.0; 2) CC-BY-SA-4.0; 3) MIT License

**Figure 1.**
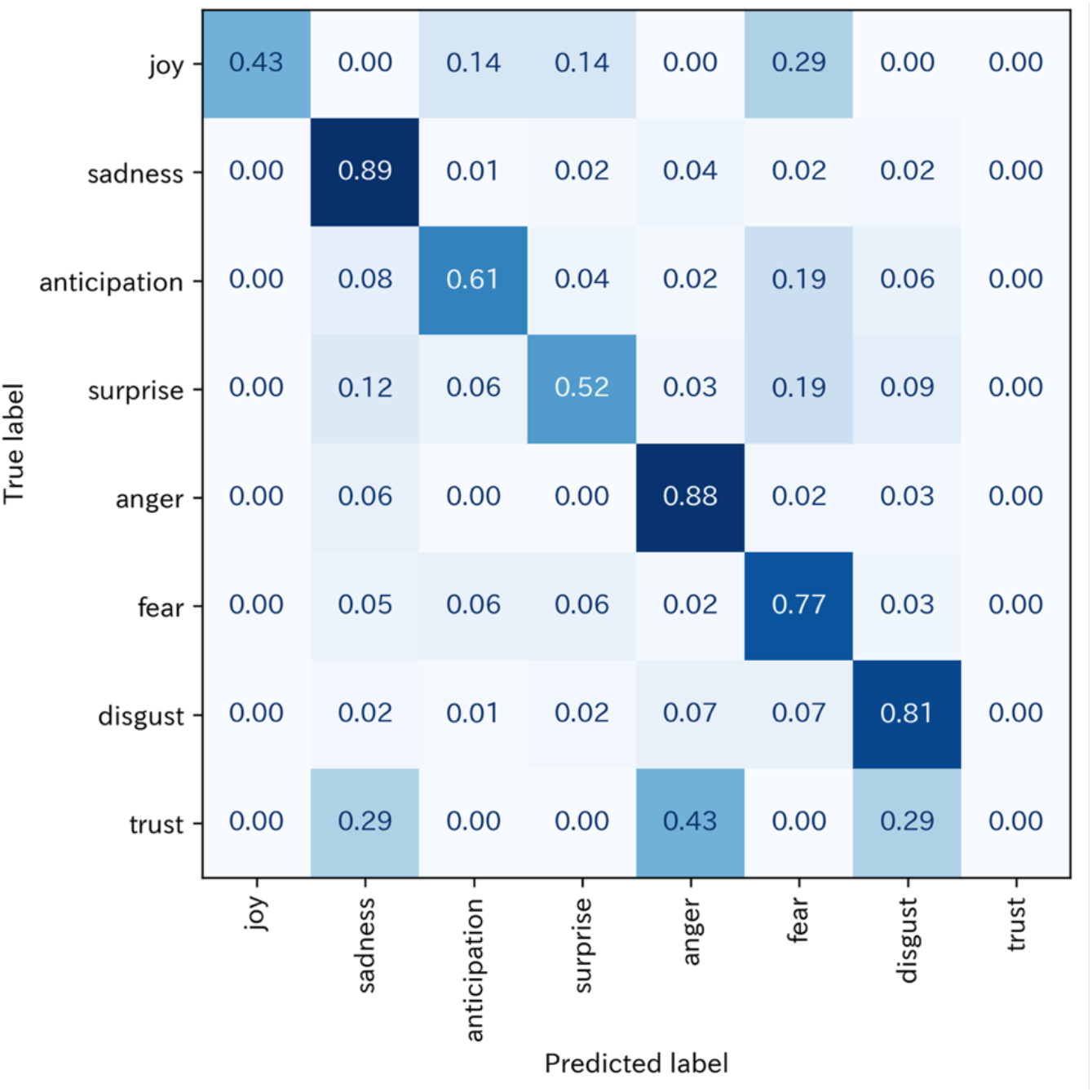
Confusion matrix of the model based on *tohoku-nlp/bert-large-japanese-v2* A confusion matrix illustrating the classification performance of the best-performing model across eight basic emotions. Values along the diagonal represent accurate classification rates, while off-diagonal elements indicate confusion rates between different emotions. Negative emotions such as anger and sadness are classified with relatively high accuracy, whereas emotions like trust are more frequently confused with other emotional states.

### 3.2. Participant Characteristics

Table 2 presents the number of participants by sex, their ages at first and final hospital visits, duration of outpatient attendance, number of visits, full-scale IQ scores, and comorbidities. In our study, although the sample size was small, comorbidities of ADHD (50%), depression (14%), and OCD (7%) were observed. These findings are consistent with previously reported rates of 45%, 14%, and 10%, respectively[14].

**Table 2.** Demographic and clinical characteristics of the participants (N=14)

|  |  |
| --- | --- |
| Sex (male/female) | 10 / 4 |
| Age at first hospital visit (years) | 12.7 (4.1) |
| Age at last hospital visit (years) | 24.9 (3.2) |
| Duration of outpatient follow-up (years) | 10.4 (2.4) |
| Number of outpatient visits (days) | 88.5 (25.0) |
| Full-scale IQ score | 91.2 (17.9) |
| Comorbidity | ADHD 50.0%, MDD 14.2%, OCD 7.0% |

Values are mean (SD).
ADHD: Attention-Deficit/Hyperactivity Disorder; MDD: Major Depressive Disorder; OCD: Obsessive-Compulsive Disorder

### 3.3. Emotion Profile

Using the best-performing model based on *tohoku-nlp/bert-large-japanese-v2*, we generated 1,239 outputs of eight-dimensional vectors representing emotion profile estimation values from the text data of the participants’ utterances in each session.

For example, when the Japanese text that translates to “That’s why I try not to take art or physical education. I don’t want to see them. Please do something. But no matter what I say, they won’t listen.” is input into the model, it generates the following output: (‘joy’: 0.063, ‘sadness’: 0.156, ‘anticipation’: 0.073, ‘surprise’: 0.076, ‘anger’: 0.102, ‘fear’: 0.120, ‘disgust’: 0.344, ‘trust’: 0.064).

As shown in Table 3, we calculated the mean and standard deviation of the probability (softmax outputs) for each of the eight emotion was across all sessions. Additionally, Table 3 displays the frequency with which each emotion was identified as predominant, measured as the number of days where that particular emotion showed the highest probability compared to other emotions.

**Table 3.** Emotion profile estimation values from BERT model analysis (n = 1,239)

|  | Mean Probability (SD) | Number of Days as<br>Predominant Emotion |
| --- | --- | --- |
| joy | 0.103 (0.116) | 154 |
| sadness | 0.266 (0.240) | 482 |
| anticipation | 0.151 (0.164) | 242 |
| surprise | 0.096 (0.096) | 77 |
| anger | 0.054 (0.047) | 3 |
| fear | 0.143 (0.117) | 146 |
| disgust | 0.136 (0.146) | 135 |
| trust | 0.051 (0.040) | 0 |

### 3.4. Principal Component Analysis

We analyzed the 1,239 emotion profile estimation values outputs using PCA, extracting four components with cumulative contribution ratios exceeding 90 %. The results for PCA and component retrieval for eight basic emotions are shown in Table 4 and 5, respectively.

**Table 4.** Summary of PCA results.

|  | PC1 | PC2 | PC3 | PC4 | PC5 | PC6 | PC7 | PC8 |
| --- | --- | --- | --- | --- | --- | --- | --- | --- |
| Eigenvalue | 0.069 | 0.030 | 0.019 | 0.015 | 0.009 | 0.003 | 0.000 | 0.000 |
| Proportion of<br>Valiance (%) | 47.1 | 20.6 | 13.0 | 10.5 | 6.5 | 2.1 | 0.2 | 0.0 |
| Cumulative<br>Proportion (%) | 47.1 | 67.7 | 80.7 | 91.2 | 97.7 | 99.8 | 100.0 | 100.0 |

**Table 5.** Component loadings for the eight basic emotions.

| Eigenvectors | PC1 | PC2 | PC3 | PC4 |
| --- | --- | --- | --- | --- |
| joy | -0.194 | -0.102 | -0.35 | -0.608 |
| sadness | 0.903 | -0.200 | 0.104 | -0.059 |
| anticipation | -0.341 | -0.666 | 0.504 | 0.209 |
| surprise | -0.116 | 0.054 | -0.281 | -0.101 |
| anger | -0.076 | 0.065 | -0.041 | -0.072 |
| fear | -0.025 | 0.145 | -0.431 | 0.749 |
| disgust | -0.074 | 0.691 | 0.582 | -0.047 |
| trust | -0.075 | 0.012 | -0.087 | -0.070 |

Notably, the first principal component (PC1), which accounting for 47.1 % of the variance, was strongly influenced by sadness (loading = 0.903). As shown in Figure 2A, the loading vector for sadness extends prominently along the positive direction of the PC1 axis, while no other emotion exhibits comparably strong loading vectors in the negative direction of PC1. This indicates that a substantial portion of variation in emotional intensity estimates is attributable to the core dimension associated with sorrowful or negatively valenced affective states.

**Figure 2.**
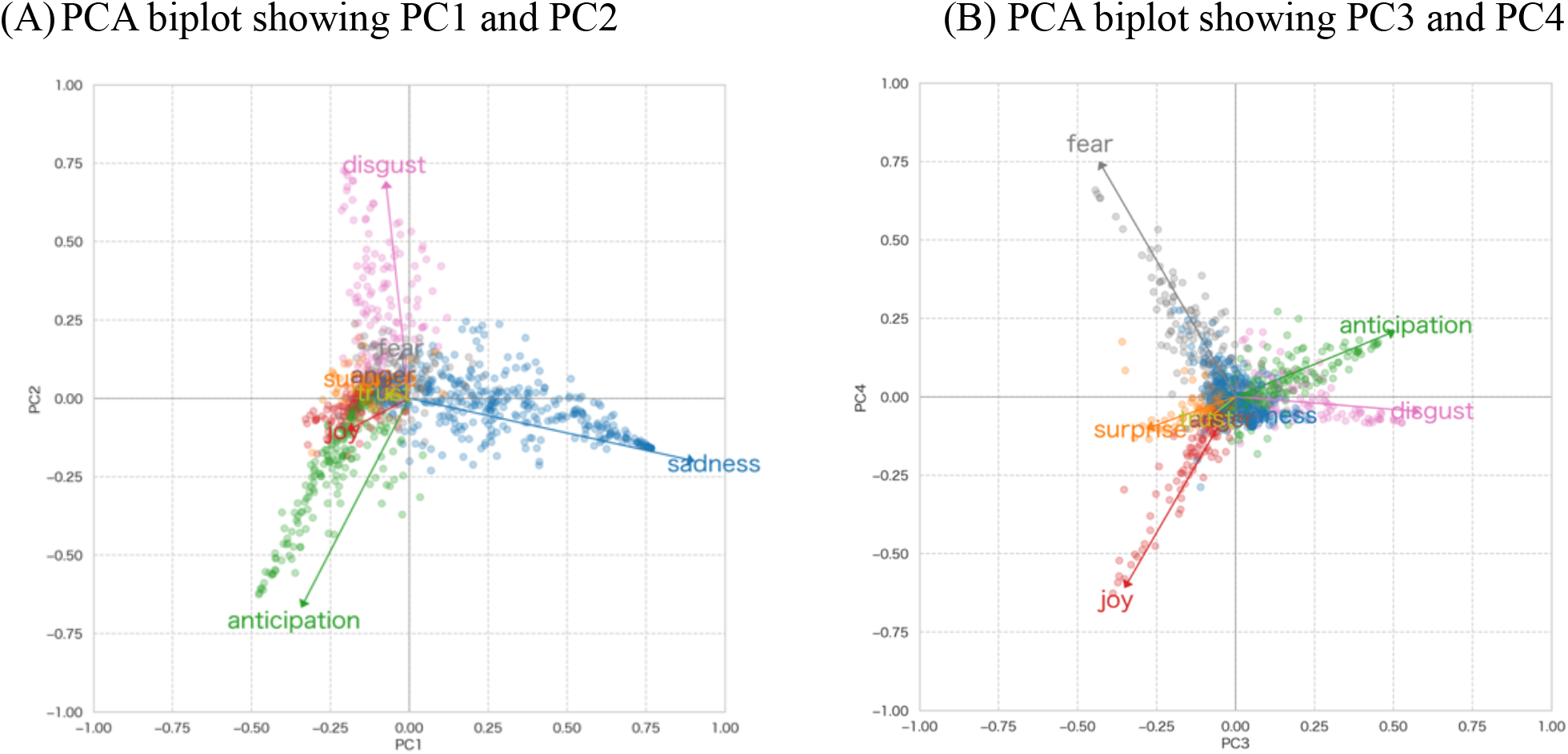
Biplot of Principal Components The biplot illustrates the relationships between variables and samples projected onto the principal components (A: PC1 and PC2; B: PC3 and PC4). Arrows represent the variable loadings, with their length and direction indicating the contribution and correlation of each variable. In these biplots, each point represents an individual session, color-coded according to the most dominant emotion detected during that session.

The second principal component (PC2), explaining 20.6 % of the variance, was characterized by a strong positive loading for disgust (0.691) and a similarly strong negative loading for anticipation (−0.666). As shown in Figure 2A, the loading vector for disgust extends in the positive direction of the PC2 axis, while the loading vector for anticipation extends in the negative direction. This component, derived from our data, revealed an axial contrast between disgust and anticipation—a pairing that does not align with the opposing pairs proposed in Plutchik’s emotion theory. According to Plutchik’s Wheel of Emotions, disgust is opposed to trust, and anticipation is opposed to surprise, forming axes within a theoretically constructed circumplex model of basic emotions.

Third and fourth principal components (PC3 and PC4) accounted for 13.0 % and 10.5 % of the total variance, respectively, which is considerably less than the first two components. Although their variance contributions are modest, we summarize them here for completeness. Within Plutchik’s dyad (combination) of emotions, the co-activation of fear and joy corresponds to the composite emotion guilt, whereas the co-activation of anticipation and disgust corresponds to cynicism. Accordingly, PC3 ranged from guilt (fear + joy) to cynicism (anticipation + disgust), whereas PC4 reflected a fear–joy polarity.

## 4. Discussion

### 4.1. Model Performance

Our study successfully fine-tuned a BERT-based model, specifically *tohoku-nlp/bert-large-japanese-v2*, to estimate emotion profile in clinical records of adolescents with ASD. The model achieved an accuracy of 78.9% and an average cosine similarity of 94.1%, demonstrating robust performance in capturing emotional nuances in clinical text.

The WRIME dataset was originally designed to incorporate emotional intensity for estimation. However, this task has proven challenging due to the large number of samples with an emotional intensity of 0. Previous studies have reported that readers struggle to identify the writer’s emotions of ‘anger’ and ‘trust’[10]. Similarly, in our model, the number of sentences with the highest estimated emotional intensity was extremely small for ‘anger’ and ‘trust’ compared to other emotions. Furthermore, when classifying sentences based on the emotion with the highest estimated intensity, the fine-tuned model achieved an accuracy of 0% for ‘trust’ (Figure 1). To address these challenges, this study reframed the task as estimating an 8-dimensional emotion vector to represent complex emotions, rather than treating it as a classification task. Consequently, the model’s performance was evaluated using cosine similarity.

### 4.2. Sadness Axis (PC1)

PCA of the eight-emotion probability vectors showed that PC1 accounted for 47.1% of the total variance. PC1 was characterized by a strong positive loading for sadness (0.903). This interpretation is consistent with longitudinal evidence that depressive symptoms peak in early to mid-adolescence among autistic youth [15] and with the dysphoria factor identified in the Emotion Dysregulation Inventory, where sadness constitutes a key but not solitary component of emotion dysregulation[16]. Ghaziuddin et al. [17]similarly highlighted depression as the most prevalent psychiatric comorbidity in autistic individuals. Taken together, our data suggest that sadness-related affect represents a principal axis of emotional variability in adolescents with ASD, although its expression is likely modulated by clinical context and comorbid depressive pathology.

All participants in the present study were autistic individuals without co-occurring intellectual disability (ID). This sampling characteristic is clinically relevant, because large cohort studies demonstrate that the risk of depressive disorders is considerably higher in autistic individuals without ID than in those with ID (adjusted RR ≈ 4.3 vs 1.8)[18]. Systematic reviews of child and adolescent samples corroborate this pattern, reporting pooled prevalence estimates of major depressive episodes ranging from 20% to 34% in non-ID groups[19]. During the transition from early adolescence to young adulthood, these individuals often develop heightened insight into their social communication differences and face novel demands in secondary education, university, or employment. Emerging evidence suggests that this combination of increased self-awareness, “social camouflaging,” and greater environmental complexity erodes self-esteem and promotes internalising symptomatology[20]. The predominance of sadness on the first principal component in our data is consistent with a broader literature indicating that dysphoric affect is a central feature of the emotional landscape for autistic youth without ID.

### 4.3. Disgust-Anticipation Axis (PC2)

PC2 accounted for 20.6 % of the total variance and featured a strong positive loading for disgust (0.691) alongside an equally strong negative loading for anticipation (–0.666). This empirically derived axis juxtaposes two emotions that are not opposite in Plutchik’s circumplex model, where disgust is paired with trust and anticipation with surprise[21]. The divergence implies that autistic adolescents may organize affective experiences along dimensions that differ from canonical circumplex structures. One interpretation is that negative reactions to potential environmental change (disgust) co-vary with diminished forward-looking expectancy (anticipation), echoing the well-documented intolerance of uncertainty and preference for sameness in ASD[22, 23]. In this context, novel or unpredictable stimuli that ordinarily elicit curiosity or anticipatory excitement in neurotypical youth may instead evoke aversive responses, effectively reversing the valence of anticipation.

Plutchik’s opposition between disgust and trust is thought to arise from social-interactional appraisals, yet social motivation is often attenuated in ASD. We therefore speculate that the emotional axis uncovered here reflects a shift from social to environmental salience: when social trust is relatively muted, the dominant counterpart to disgust becomes the expectation of change itself. It is important to note that PCA loadings are influenced by both participants’ linguistic expressions and the characteristics of the emotion-classification model; replication with alternative instruments will be necessary to confirm this pattern.

### 4.4. Secondary Components (PC3/4)

The interpretability of PC3 (guilt to cynicism) and PC4 (fear–joy balance) is intriguing; however, each explains just over one-tenth of the total variance. We therefore regard these components as hypothesis-generating rather than definitive. Future studies with larger samples and longitudinal designs will be needed to determine whether these secondary axes have replicable clinical significance.

### 4.5. Clinical Implications

Although our study was limited to quantitative emotion analysis, two cautious clinical inferences emerge. First, the marked salience of sadness supports using a low threshold for depression screening in autistic adolescents, consistent with longitudinal data showing persistently elevated depressive trajectories in this group[15]. Second, the contrasted disgust–anticipation axis parallels findings of sensory-driven disgust processing and the well-established link between intolerance of uncertainty and emotional distress in autism[24, 25]. Together, these observations tentatively point to the value of sensory-modulation and uncertainty-management strategies, while underscoring the need for targeted trials before formal clinical recommendations can be made.

### 4.6. Limitations and Future Directions

Our findings should be interpreted in light of three main limitations. First, the modest sample size restricts external validity; the present results are best regarded as hypothesis-generating until they can be replicated in larger, multisite cohorts. Second, we inferred emotions from clinicians’ notes rather than from direct assessments, so the extracted signals may partly reflect individual documentation styles and interpretive biases. Third, although the classifier performed well overall, its accuracy for rarely expressed emotions—most notably trust—remains suboptimal.

These caveats suggest several priorities for future work. Longitudinal designs that follow adolescents with ASD through key developmental transitions could clarify how affective patterns evolve over time. Cross-condition comparisons (e.g., ADHD, specific learning disorders) would help delineate shared versus disorder-specific emotional signatures. Finally, linking emotion trajectories to treatment engagement and clinical outcomes could inform more precisely targeted interventions for ASD.

## 5. Conclusion

This study employed advanced NLP techniques to analyze the emotional characteristics of adolescents with ASD through their clinical records. Our findings revealed a dominant role of sadness and a unique contrast between disgust and anticipation which diverges from theoretical frameworks such as Plutchik’s Wheel of Emotions. These results underscore the potential of machine learning models to uncover nuanced emotional dynamics in ASD and offer valuable insights for clinicians and researchers to tailor interventions for this population.

## Data Availability

All data produced in the present study are available upon reasonable request to the authors.

https://huggingface.co/MuneK/bert-large-japanese-v2-finetuned-wrime

## Funding

No funding was received for conducting this study. The authors have no relevant financial or non-financial interests to disclose.

## Notes

### Competing Interest Statement

The authors have declared no competing interest.

### Funding Statement

This study did not receive any funding.

### Author Declarations

the Ethical Review Committee of Yamagata University Faculty of Medicine gave ethical approval for this work.

